# Integrated Analysis of the Association Between Alzheimer’s Disease (AD) and Cardiovascular Disease (CVD)

**DOI:** 10.1101/2021.06.15.21258992

**Authors:** Katherine Wei, Anna Delprato

## Abstract

Alzheimer’s Disease (AD), the sixth leading cause of death in the US, and cardiovascular disease (CVD), the first leading cause of death in the US, are frequently associated. Past studies hypothesize that amyloid deposits damage organs, causing this association. Examining how vascular factors can influence AD pathogenesis can help in understanding the link between the blood to the brain, which can provide alternative paths of exploration for disease treatment. This study analyzes gene expression and shared biological processes between AD and CVD, specifically myocardial infarction and heart failure, via bioinformatic approaches and published datasets from the Gene Expression Omnibus (GEO). First, 73 differentially expressed genes (DEGs) were identified among four datasets using blood samples from AD and CVD patients. Panther’s Gene Ontology Analysis validated several biological processes such as xylulose biosynthetic process and toll-like receptor TLR1:TLR2 signaling pathway along with molecular functions, cellular components, and pathways to be significantly enriched in the list of 73 DEGs. Analysis of protein-protein interactions and the associated gene network indicated that from the list of 73 DEGs, only six (MAPK14, TLR2, HCK, GRB2, PRKCD, PTPN6) had eight or more degrees. Next, those six genes were identified in a normalized dataset containing different brain regions of AD and non-AD patients. Two-sample t-tests for differences in mean showed statistically significant differences in GRB2 and PRKCD, supporting a blood-brain relationship in the association between AD and CVD. This study can help in developing new medications to target AD and CVD susceptible genes.

## Introduction

### Alzheimer’s Disease (AD) Background

Over 5 million people live with AD in the U.S. in 2020 making it the 6th leading cause of death in the U.S. Even though the rate of mortality with AD is higher than that of breast cancer and prostate cancer combined, the amount of AD research is minimal compared to that of cancer research and other diseases [1]. The symptoms associated with AD are memory loss, confusion, difficulty completing familiar tasks, and inability to understand basic images and words [2]. Current researchers have discovered that the accumulation of proteins like beta-amyloid and tau in neural tissue has been the root cause of the disease. There are many genes associated with AD such as the amyloid precursor protein (APP), apolipoprotein (APOE), and phospholipase D3 (PLD3) [3].

Studies conducted with pluripotent stem cells and clustered regularly interspaced short palindromic repeats (CRISPR) have showcased a strong correlation between some genes and late-onset AD [4]. For instance, the apolipoprotein (APOE) gene, which functions in the transport of brain cholesterol and promotion of lipoprotein clearance from circulation, has been heavily studied recently. It has three alleles: E2, E3, and E4. The E2 allele is considered protective and has a worldwide frequency of 4.2%, while E3 is the most common allele with a frequency of 77.9%. Finally, the E4 allele has been found to be the strongest risk factor associated with AD with a 13.7% worldwide frequency but a ∼40% frequency with patients who have AD. Since identifying the risk of the development of AD-related to APOE4, researchers have studied a lot about the gene’s function, structure, and sequence. All three alleles have one or two different amino acid substitutions. For instance, between E3 and E4, the amino acid difference lies in position 112 where E3 has cysteine and E4 has arginine [5]. Infants that carry the APOE4 gene have been found to have less gray matter than normal infants. Less gray matter typically translates to limited communication between neurons and other cells [6].

### Cardiovascular Disease (CVD) Background

CVD or heart disease is the leading cause of death in the U.S. with about 655,000 people dying from CVD every year, costing roughly $219 billion in terms of healthcare services, medicines, and lost productivity due to death. CVD includes coronary heart disease (CAD), heart attack (also called myocardial infarction), heart failure, stroke, atherosclerosis, and others [7]. For many of these diseases, DNA inheritance plays a role in its pathogenesis. Many studies have established this, for instance, Lloyd-Jones et al concluded that parents who have CVD increase the risk of it inheriting to their offspring by 3 times [8].

The two primary types of cardiovascular disease that this study focuses on are myocardial infarction (MI) and heart failure. MI occurs when blood flow to the heart is blocked either from buildup of fat, cholesterol, or other substances in coronary arteries [9]. Every year in the U.S. about 805,000 Americans have heart attacks [7]. Heart failure occurs when heart muscles don’t pump as much blood as needed [10]. About 6.2 million adults in the U.S. have heart failure [11]. Several CVDs have common risk factors such as age, obesity, tobacco and drug usage, and stress. The most notable risk factor in relation to this study is family history.

One significant gene studied in the past and has shown a link with CVD is low-density lipoprotein (LDL), which is a major cholesterol-carrying lipoprotein [12]. A deficit of LDL receptors and transport promotes cholesterol absorption and LDL synthesis, thus increasing the onset of vascular pathologies. In a study focused on MI, Yamada et. al. found a statistically significant association of specific single-nucleotide polymorphisms (SNPs) in the connexin 37 gene in men and the plasminogen-activator inhibitor type 1 gene and the stromelysin-1 gene in women. The SNPs analyzed may signal a correlation of susceptibility to MI in those patients [13]. Another study on congestive heart failure in African Americans suggests that the presence of both the α2c-adrenergic receptor, associated with norepinephrine release at cardiac sympathetic-nerve synapses, and β1-adrenergic receptor, associated with increased sensitivity of cardiomyocytes to norepinephrine, increase heart failure risk [14].

### CVD and AD Association

Past studies have demonstrated a significant link between CVD and AD. For instance, research with AD autopsies showed an association between microvascular ischemic lesions and Alzheimer lesions [15]. Another relevant association that may indicate a relationship between CVD and AD is the one between carotid artery wall thickness, ankle-arm index, and AD which was studied in the prospective Cardiovascular Health Study [16]. This may be largely due to the hypothesis that the underlying mechanisms are hypoperfusion and emboli. This association between CVD and AD is also possibly related to the fact that both diseases involve amyloid deposits [17]. Other studies regarding the relationship between CVD and AD hypothesize that the connection maybe due to the common risk factors, or that CVD indirectly damages the brain and thus influences neurodegeneration, or that CVD directly affects AD development due to neuron death or accumulation of amyloid plaques, similar to the hypothesis of the first study mentioned [18]. More recently, beta-amyloid aggregations, a strong indicator of AD, have been found in the hearts of patients with idiopathic dilated cardiomyopathy. This finding further encourages a genomic analysis for the link between CVD and AD [19]. Because both CVD and AD are linked with genetics, an integrated analysis can help to further explore the connections between the brain and heart by studying the correlation between CVD and AD.

### Research Goals

The goal of this project is to identify the biological processes, molecular functions, and cellular components of common genes among AD and CVD to understand why these diseases overlap in many patients. Another goal is to understand the relationship between AD and CVD to allow for future developments towards treating AD by managing CVD.

## Materials and Methods

### Bioinformatics Resources Used

This study primarily consists of bioinformatic approaches using published datasets, software, and tools. To comprehensively analyze the association between AD and CVD, two phases of research were conducted: 1) in the blood and 2) in the brain. The data for this study were retrieved from the National Center for Biotechnology Information’s (NCBI) Gene Expression Omnibus (GEO) database, which contains gene expression datasets published for the public to use. GEO’s analytical tool GEO2R was used to obtain quantitative data to filter out statistically significant genes to be further analyzed in the study. Microsoft Excel was used to screen out non-statistically significant genes and sort overexpressed and underexpressed genes together in sheets. The Search Tool for the Retrieval of Interacting Proteins (STRING), a database of known and predicted protein-protein interactions obtained from genomic context predictions, high throughput lab experiments, co-expression, automated textmining, and previous knowledge in database, was used in order to better understand the relationships between genes and analyze their active roles on the risk of CVD and AD. STRING also calculates statistically significant Gene Ontology processes and pathways. Cytoscape, a software for visualizing complex networks, was used to obtain specific data on the STRING network (degrees, clustering coefficient). The gene ontology (GO) resource powered by Panther displayed the false discovery rate (FDR), a measure of how many expected false discoveries there are (rate of type I errors), and significant biological processes, allowing for better analysis. The SOURCE tool, a search tool created from the compilation and collection of genomic databases like UniGene, Swiss-Prot, OMIM, and Entrez Gene that captures the genetics and molecular biology of genes of several organisms into a GeneReport, was used to obtain detailed information on individual genes.

All tables were generated by Excel. Figures illustrating protein-protein interactions were obtained from String DB. To identify the common genes among different datasets, Venny 2.1, an interactive tool for comparing lists with Venn diagrams powered by Bioinformatics for Genomics and Proteomics (BioinfoGP), was used. JMP Pro 12, a software for data analysis, was used to obtain powerful statistical analysis of the brain dataset. The graphs and figures provided by JMP Pro 12 of the brain dataset analysis were copied into this paper.

For gene query, the GEO dataset browser’s data analysis tool to find genes was used. After the genes of interest are identified from a specific dataset, the graph and gene expression table for the experimental and control units from the dataset were retrieved to analyze. The data was then divided based on brain region (hippocampus, frontal cortex, temporal cortex) and AD or non-AD patients. Afterward, a 2-sample t-test for difference in mean was conducted on JMP Pro 12 to determine statistically significant differentially expressed genes in the brain regions.

### Data Collection

For analysis of the blood, a total of seven datasets, four CVD and three AD datasets, were retrieved from the GEO database by using search terms like “Alzheimer’s Disease,” “dementia” combined with “cardiovascular disease,” “heart disease,” “coronary artery disease,” “myocardial infarction,” and “heart failure.” All datasets consist of only homo sapiens, blood samples, expression profiling by array, and display a normal distribution without outliers and/or have more than 30 samples.

The two datasets chosen for analysis of AD blood were ai) GSE63060, aii) GSE63061. Dataset ai) which was submitted November 6, 2014, and updated on May 3, 2019, is by Angela Hodges and Robert Howard. This dataset contains 329 samples: 145 AD patients, 80 mild cognitive impairment (MCI) patients, and 104 age and gender-matched controls. For consistency, the MCI patients were omitted from the analysis. Dataset aii) is also by Angela Hodges and Robert Howard, submitted November 6, 2014, and updated on May 3, 2019, since this is batch two of their study. This dataset contains 388 samples: 139 AD patients, 109 MCI patients, 3 borderline MCI patients, 134 controls, and 3 miscellaneous samples. For consistency, only the AD and controls were used in this analysis.

**Table 1.** Dataset information for GSE63060, GSE63061, GSE48060, and GSE9128. In datasets GSE63060 and GSE6306, the control and experimental samples don’t add up to the total samples because they included mild cognitive impairment patients that were omitted from this study.

| Series Accession | Disease of Interest | Control | Experimental | Author | Total Samples |
| --- | --- | --- | --- | --- | --- |
| GSE63060 | Alzheimer's Disease | 104 | 145 | Hodges A, Howard R | 329 |
| GSE63061 | Alzheimer's Disease | 134 | 139 | Hodges A, Howard R | 388 |
| GSE48060 | Myocardial Infarction | 21 | 31 | Suresh R, Li X, Chiriac A, Goel K, Terzic A, Perez-Terzic C, Nelson TJ | 52 |
| GSE9128 | Heart Failure | 3 | 8 | Napolitano M, Capogrossi MC | 11 |

The two datasets chosen for analysis of CVD blood were bi) GSE48060, and bii) GSE9128. Dataset bi) which was submitted June 18, 2013, and updated March 25, 2019, is by Rahul Suresh et. al. This dataset contains 52 samples: 21 control samples and 31 experimental MI samples. Dataset bii) which was submitted September 21, 2007, and updated August 10, 2018, is by Monica Napolitano and Maurizio C Capogrossi. This dataset contains 11 samples: 3 control samples and 8 experimental heart failure samples.

After the datasets were identified, GEO’s analytical tool GEO2R was used to define the groups in each dataset into control and experimental, ensuring that the experimental groups were defined first then the control so that the logFC numbers were accurate. Afterward, any genes that had a p-value > 0.05 were omitted from the study because they weren’t statistically significant. Afterward, the genes that were statistically significant were sorted into overexpressed and underexpressed genes based on logFC values. Genes with logFC values > 0 were classified as overexpressed and genes with logFC values < 0 were classified as underexpressed. After the underexpressed and overexpressed genes were labeled, underexpressed genes in all 4 datasets (GSE63060, GSE63061, GSE48060, GSE9128) were compared to identify the common underexpressed genes and the same process was done with the overexpressed genes.

The list of the common 73 underexpressed and overexpressed genes was entered into STRING which was able to identify all 73 of the inputted genes. STRING constructed a network of the protein-protein interactions of the genes. The settings were changed so that disconnected nodes were hidden from the network in order to improve clarity. The minimum required interaction score was medium confidence (0.400).

The STRING network was then sent to Cytoscape for deeper analysis. The Network Analyzer tool in Cytoscape calculated metrics such as node degree and clustering coefficient that STRING did not have. Genes that had a degree (the number of genes connected to a certain gene) of eight or larger were considered genes of interest as they showed significant interactions with other genes. The list of the 73 underexpressed and overexpressed genes was then entered into the Gene Ontology (GO) enrichment analysis. The settings of GO were set to GO biological process complete, Fisher’s Exact, and Calculate False Discovery Rate (FDR). A list of biological processes and pie charts illustrating the details were retrieved for further analysis.

For analysis of the brain, the dataset chosen was GSE36980 titled “Expression data by post mortem Alzheimer’s Disease brains” by Yusaku Nakabeppu et. al. which was submitted April 2, 2012, and updated September 6, 2020. The study used an interspecies comparative microarray analysis using RNAs prepared from postmortem human brain tissues donated for the Hisayama study, which was a population-based prospective cohort study [20]. This dataset contains 80 samples: 15 samples of frontal cortex (FC) with AD, 18 samples of FC non-AD, 8 samples of hippocampus (HI) with AD, 10 samples of HI non-AD, 10 samples of temporal cortex (TC) AD, and 19 samples TC non-AD. All brain regions originally contained 33,298 genes, but after the non-statistically significant data was filtered out, there were only 8441 in the hippocampus, 3828 in the frontal cortex, and 4834 in the temporal cortex.

## Results

### Blood Analysis

After filtering out the non-statistically significant genes, there were a total of 73 shared genes (Appendix) from the four datasets (GSE63060, GSE63061, GSE48060, GSE9128) used to analyze blood from AD and CVD patients. The list of 73 shared genes among all 4 datasets consisted of 31 shared underexpressed genes and 42 shared overexpressed genes. There were also 367 underexpressed genes and 432 overexpressed genes shared in at least 3 datasets. For clarity, only the list of 73 shared genes in all 4 datasets was studied in this blood analysis.

The STRING analysis of the final gene list showed 84 edges in comparison to the expected 50 edges, which meant that the network had significantly more interactions than expected for a random set of genes of similar size. This signifies that these genes are likely to be partially connected as a group. Functional enrichment in the network showed xylulose biosynthetic process, cellular response to triacyl bacterial lipopeptide, and toll-like receptor TLR1:TLR2 signaling pathway as the top three out of 159 biological processes. The top three enriched molecular functions out of twelve in the network were transferase activity, scavenger receptor binding, and N-formyl peptide receptor activity. The top three cellular components enriched out of 22 were ficolin-1-rich granule membrane, ficolin-1-rich granule, and ficolin-1-rich granule lumen.

**Table 2.**
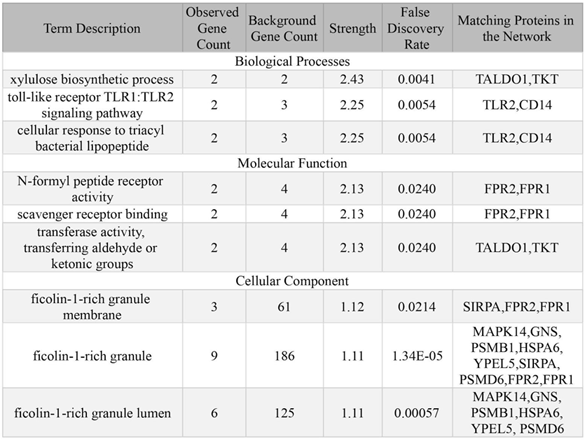
Functional enrichments in biological processes, molecular function, and cellular components retrieved from STRING ranked by strength, which is measured by log10 (observed/expected), showing how large the enrichment factor is.

The network was then sent to Cytoscape which calculated the degree (the number of genes connected to the gene of interest), clustering coefficient (a measure of how clustered nodes are around a single node with 1 being the densest and 0 being least dense), topological coefficient (a measure of how many shared neighbors are around a node with more than 1 neighbor), and expression of the gene in different tissues such as blood, heart, and nervous system [21]. Cytoscape found that the MAPK14 gene was the most connected node in the network with 14 degrees. From a SOURCE search, we found that MAPK14 (mitogen-activated protein kinase 14) is involved in proliferation, differentiation, transcription regulation, and development and activated by various environmental stresses and proinflammatory cytokines. MAPK14 was overexpressed in all datasets.

**Table 3.** Analysis from Cytoscape of the genes of interest ranked by degree.

| Gene of Interest | Blood Tissue | Heart Tissue | Nervous System Tissue | Clustering Coefficient | Topological Coefficient | Degree |
| --- | --- | --- | --- | --- | --- | --- |
| MAPK14 | 4.623546 | 2.990975 | 4.460733 | 0.1538461538 | 0.1942857143 | 14 |
| TLR2 | 4.745894 | 3.101069 | 2.934081 | 0.3111111111 | 0.2636363636 | 10 |
| HCK | 3.606493 | 2.126198 | 4.459929 | 0.4418604651 | 0.28 | 9 |
| GRB2 | 4.643225 | 3.082949 | 4.974375 | 0.2777777778 | 0.2393162393 | 9 |
| PRKCD | 4.634836 | 2.684684 | 4.568752 | 0.25 | 0.2608695652 | 8 |
| PTPN6 | 4.807801 | 1.899531 | 2.404745 | 0.1071428571 | 0.2142857143 | 8 |

The gene list was then entered into Gene Ontology (GO) enrichment analysis (powered by Panther), which identified statistically significant biological processes, including 9 processes that had a fold enrichment >100: xylulose biosynthetic process, cellular response to triacyl bacterial lipopeptide, response to triacyl bacterial lipopeptide, pentose biosynthetic process, cellular response to diacyl bacterial lipopeptide, response to diacyl bacterial lipopeptide, toll-like receptor TLR6:TLR2 signaling pathway, toll-like receptor TLR1:TLR2 signaling pathway, and xylulose metabolic process. Of these biological processes, the xylulose biosynthetic process had the smallest p-value (7.76E-05) and smallest FDR (1.85E-02). Enrichment is calculated by taking the number of genes in the list that belong to a specific biological process and dividing them by the expected number of genes that would belong if taken from a random sample of genes. A higher enrichment value indicates that a greater amount of genes in a list are involved in a certain GO process. Cellular response to bacterial lipopeptide, response to bacterial lipopeptide, and pentose-phosphate shunt, non-oxidative branch were also enriched.

The enriched biological processes shown by GO were largely in agreement with what was found by STRING: xylulose biosynthetic process, toll-like receptor TLR1:TLR2, and cellular response to triacyl bacterial lipopeptide. Panther showed no statistically significant enriched molecular functions; however, it did show 15 enriched cellular components, which were also largely in agreement with what was found by STRING with ficolin-1-rich granule and ficolin-1-rich granule lumen having a fold enrichment over 13. Panther GO enrichment analysis also showed enriched pathways, which is helpful in understanding what the genes are involved in in the body. The most enriched reactome pathways were Insulin effects increased synthesis of Xylulose-5-Phosphate, Formyl peptide receptors bind formyl peptides and many other ligands, MyD88 deficiency (TLR2/4). The most enriched panther pathways were Pentose phosphate pathway, B cell activation, Ras Pathway.

**Table 4.**
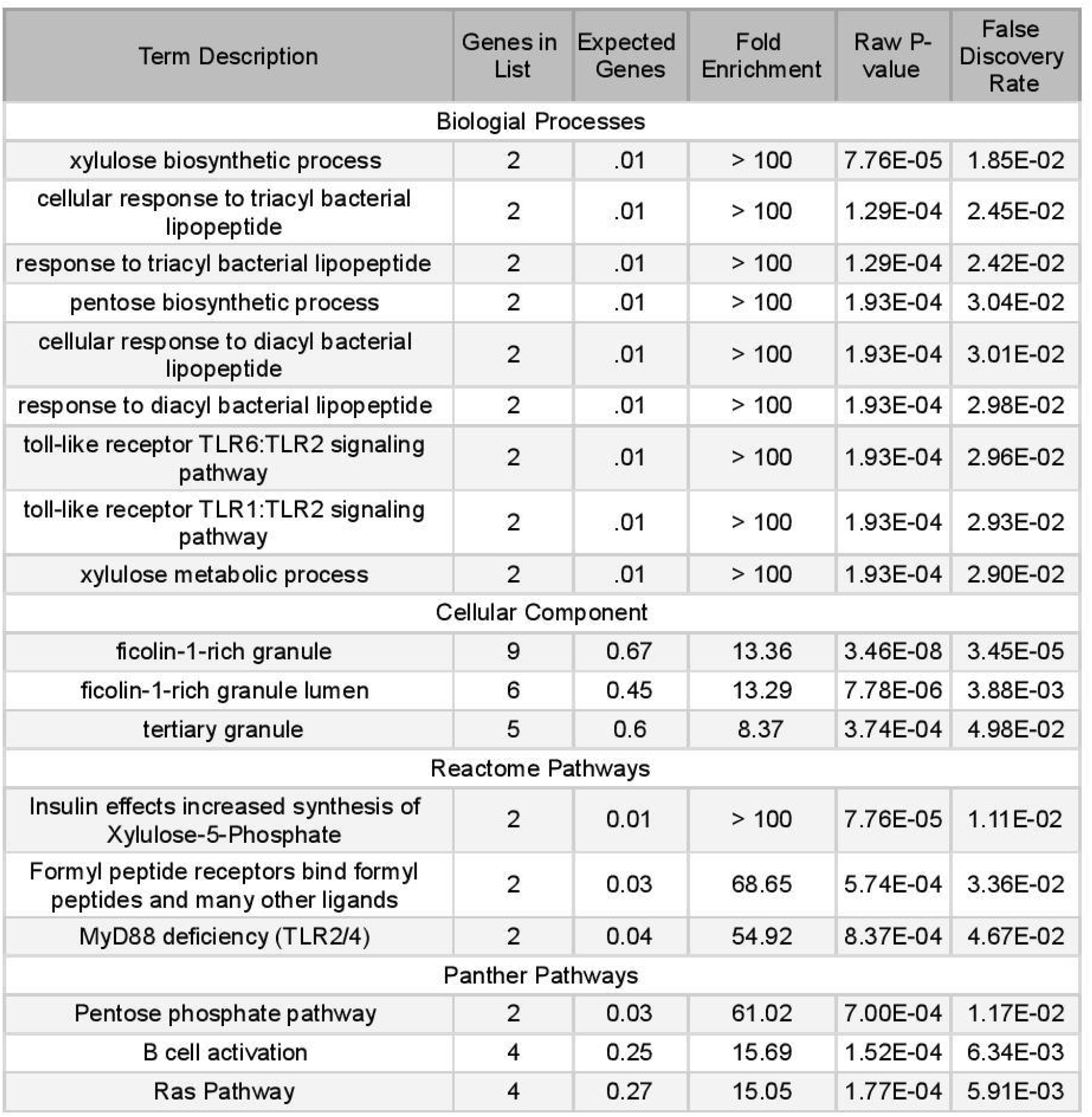
Panther Classification System’s GO enrichment analysis of various biological processes, cellular components, rectome and panther pathways ranked by fold enrichment.

### Brain Analysis

Six genes of interest identified by Cytoscape (MAPK14, TLR2, HCK, GRB2, PRKCD, PTPN6), and the associated data were extracted in the dataset GSE36980 using NCBI’s dataset browser and data analysis tools. The gene expression levels displayed in the tables provided in the browser were copied into Excel and were divided into six groups within each gene (AD HI, non-AD HI, AD FC, non-AD FC, AD TC, non-AD TC) in order to examine the pair differences between AD and non-AD by brain region. The mean and standard deviation for all pair groups within each gene were calculated using 2-sample t-tests for difference in mean between AD and non-AD units. The null hypothesis was that there was no difference between the AD mean and the non-AD mean. The alternate hypothesis was that there was a significant difference between the AD mean and the non-AD mean. If the p-value was less than 0.05, then the result was statistically significant. The genes that passed the t-test were GRB2 and PRKCD.

Firstly, the raw data from each group of GRB2 and PRKCD are imported into JMP for overall variability analysis shown as in Figure 4. It indicates that the overall gene expression of GRB2 is stronger than that of PRKCD. The gene expression for GRB2 shows FC< HI < TC for each pair of AD and non-AD. The FC<HI<TC trend is seen within the PRKCD gene as well. AD_HI is significantly smaller than non-AD_HI for PRKCD. AD is lower than non-AD for all groups (FC, HI, and TC) and for both genes.

**Figure 1.**
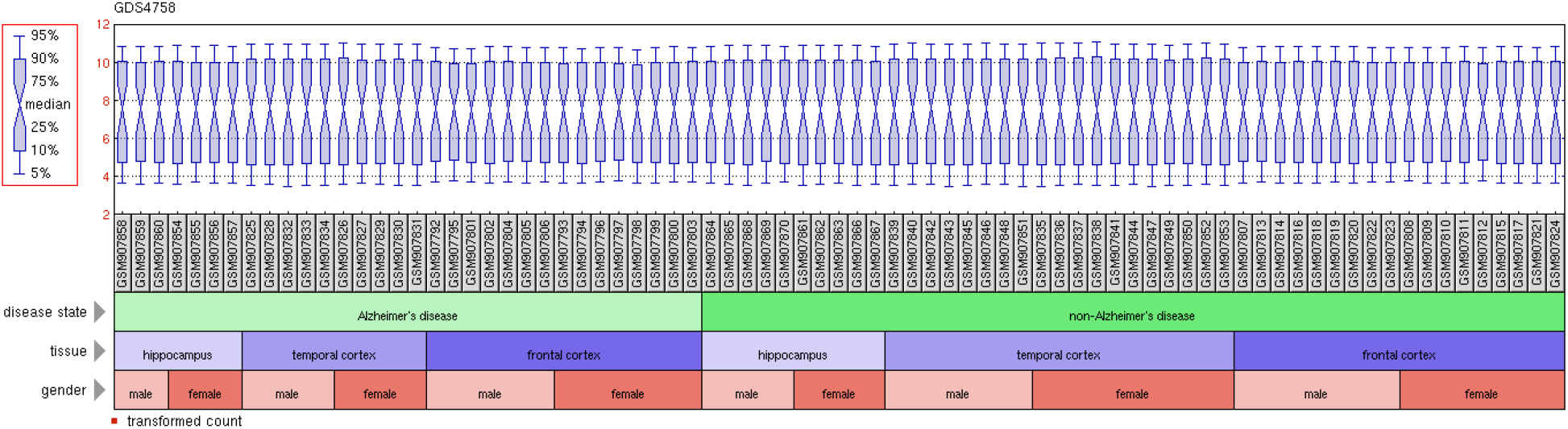
Boxplot detailing dataset analysis of GSE36980 that shows a normal distribution with no strong outliers or skewness. Figure retrieved from GEO DataSet Browser’s data analysis tool for experiment design and value distribution.

**Figure 2.**
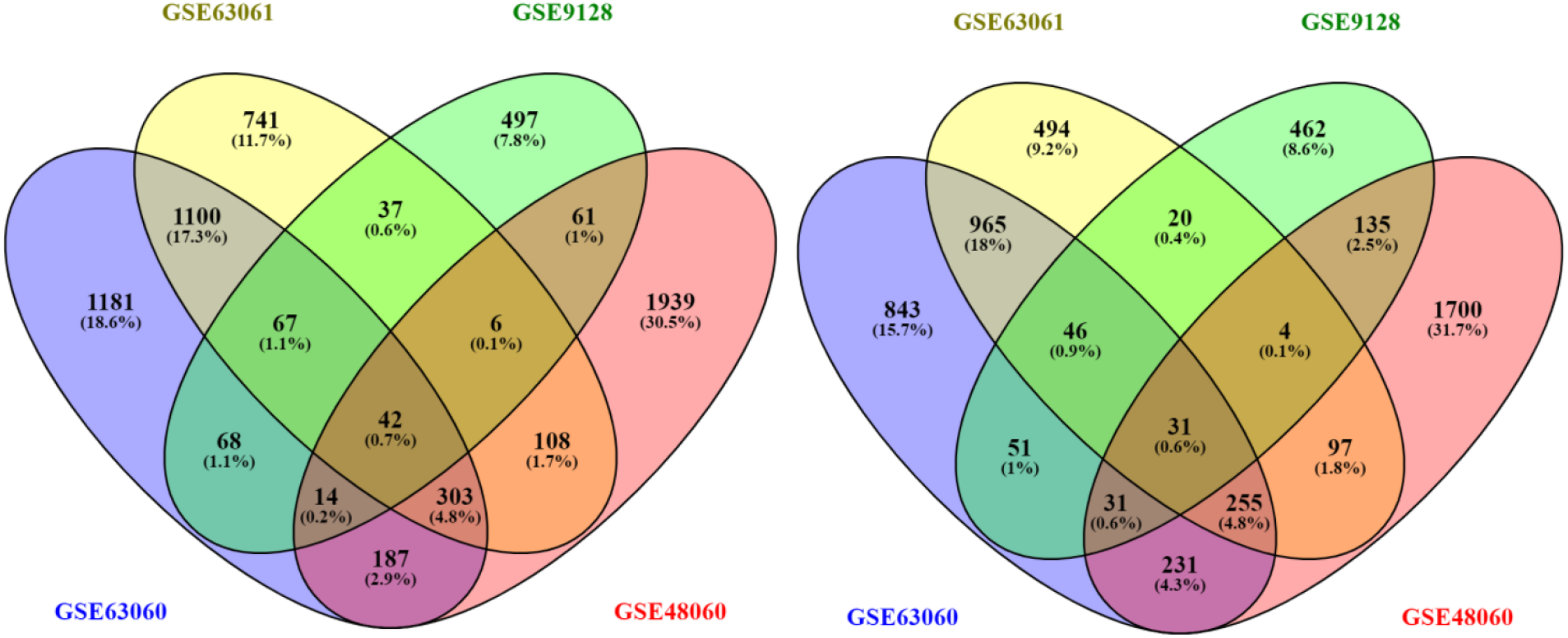
Venn diagrams of common genes in AD and CVD made by Venny 2.1. i) on the left shows the 42 common overexpressed genes in all 4 datasets. ii) on the right shows the 31 common underexpressed genes in all 4 datasets.

**Figure 3.**
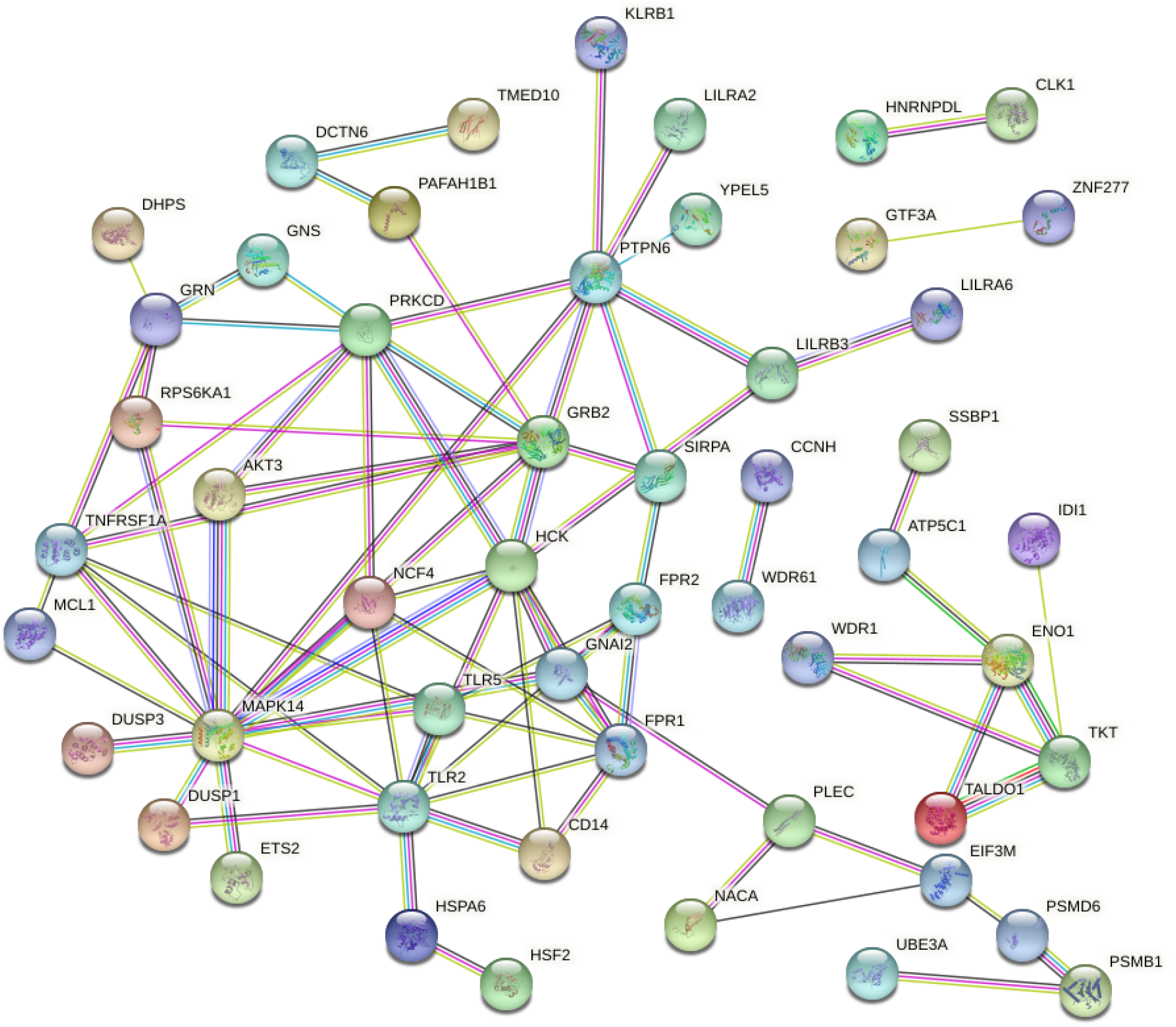
Diagram of the protein interactions between 73 differentially expressed genes retrieved from STRING. The nodes represent proteins and the edges represent the protein-protein interaction. The color of the nodes has no significant meaning but the colors of the edges represent how the interaction was determined (for example, blue is from curated databases, and pink is experimentally determined). The bottom left of the diagram shows a concentration of interactions, especially around MAPK14. Disconnected nodes, or genes with no interactions, were removed from the diagram for clarity.

**Figure 4.**
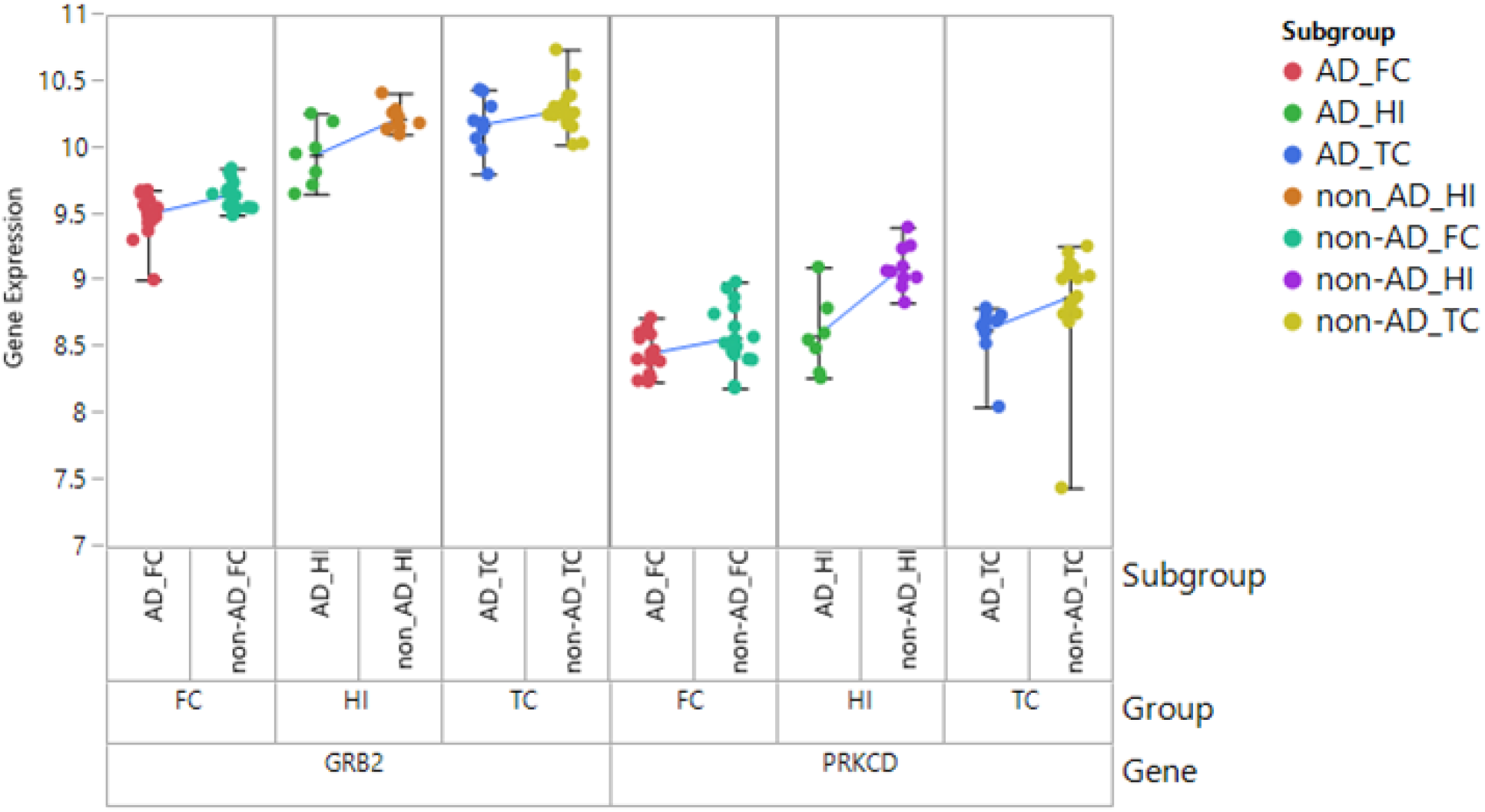
Figure showing the individual data points for the six groups, which were compared by first gene (GRB2, PRKCD), then brain region (FC, HI, TC), then the experimental group (AD, non-AD). The blue line compares the experimental pairs by mean.

Secondly, the means, standard deviation, standard error mean, lower 95% of the data, and upper 95% of the data along with the two-sample t-tests, such as p-value, degrees of freedom (df), test statistic, difference, and density curve for each subgroup are calculated for further analysis. As shown in Figure 5 to Figure 7, there is a statistically significant difference for FC and HI subgroups between AD and non-AD pairs, which is consistent for GRB2 and PRKCD genes. The TC also shows a statistically significant difference in PRKCD but not GRB2.

**Figure 5.**
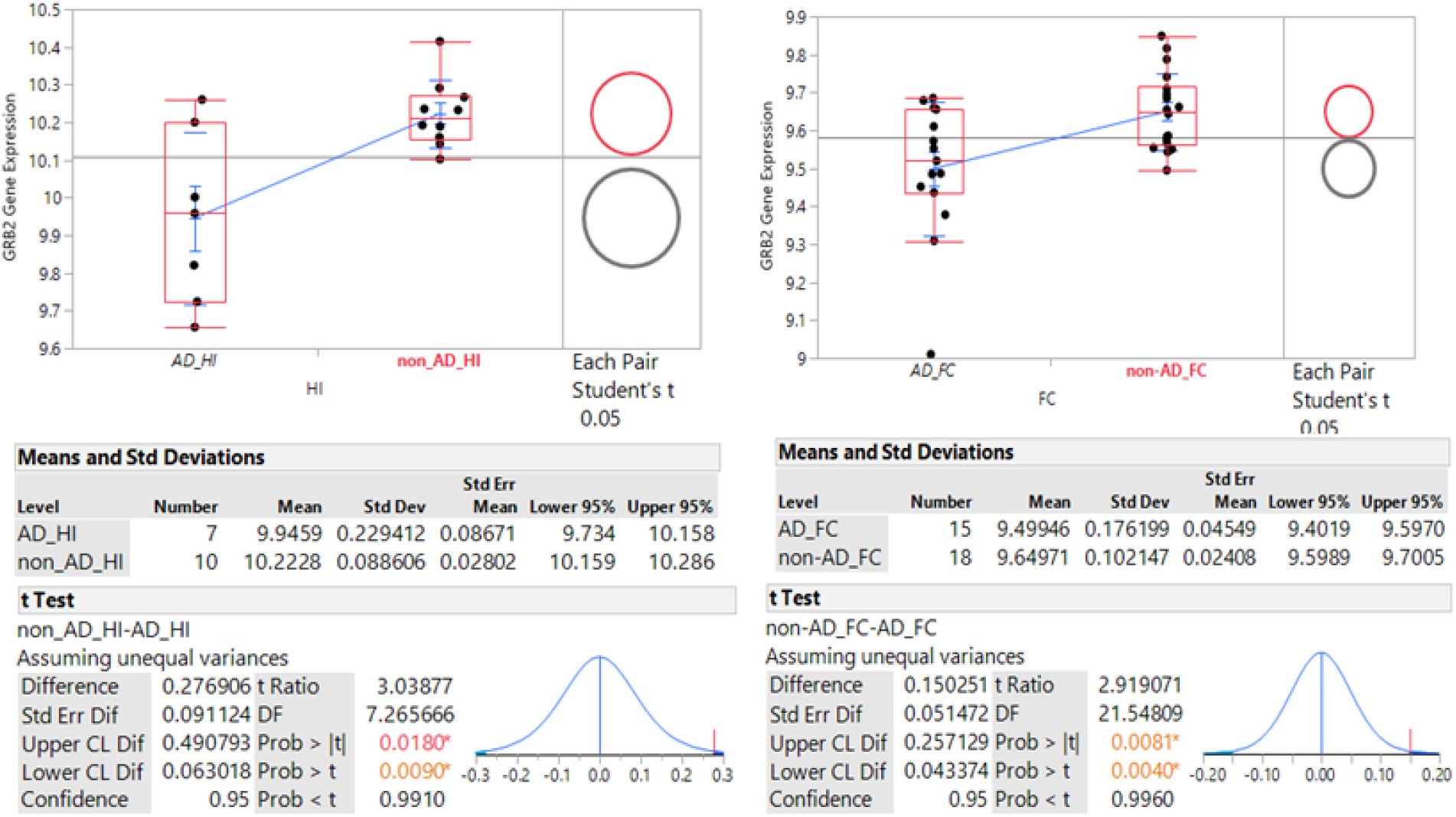
GRB2 Gene Expression in a) on the left in the HI and b) on the right in the FC. P<0.05 indicates a significant difference between AD and non-AD pairs for HI and FC subgroups. Upper CL Dif and Lower CL Dif give the 95% CI for the true difference. Note that the difference in the HI is visibly larger than the FC.

**Figure 6.**
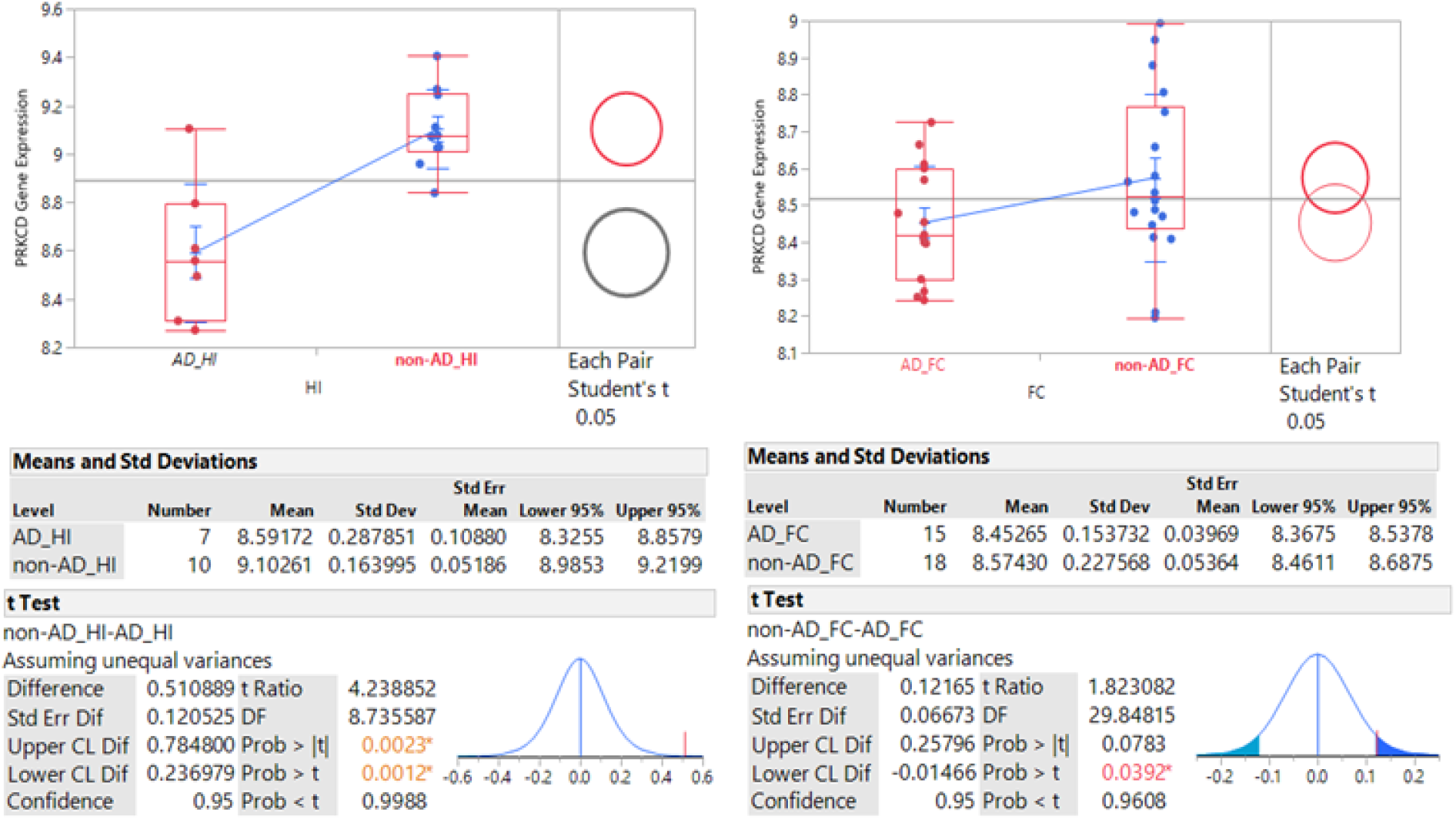
PRKCD Gene Expression a) on the left in the HI and b) on the right in the FC. Upper CL Dif and Lower CL Dif give the 95% CI for the true difference. Note that the difference in the HI is visibly larger than the FC.

**Figure 7.**
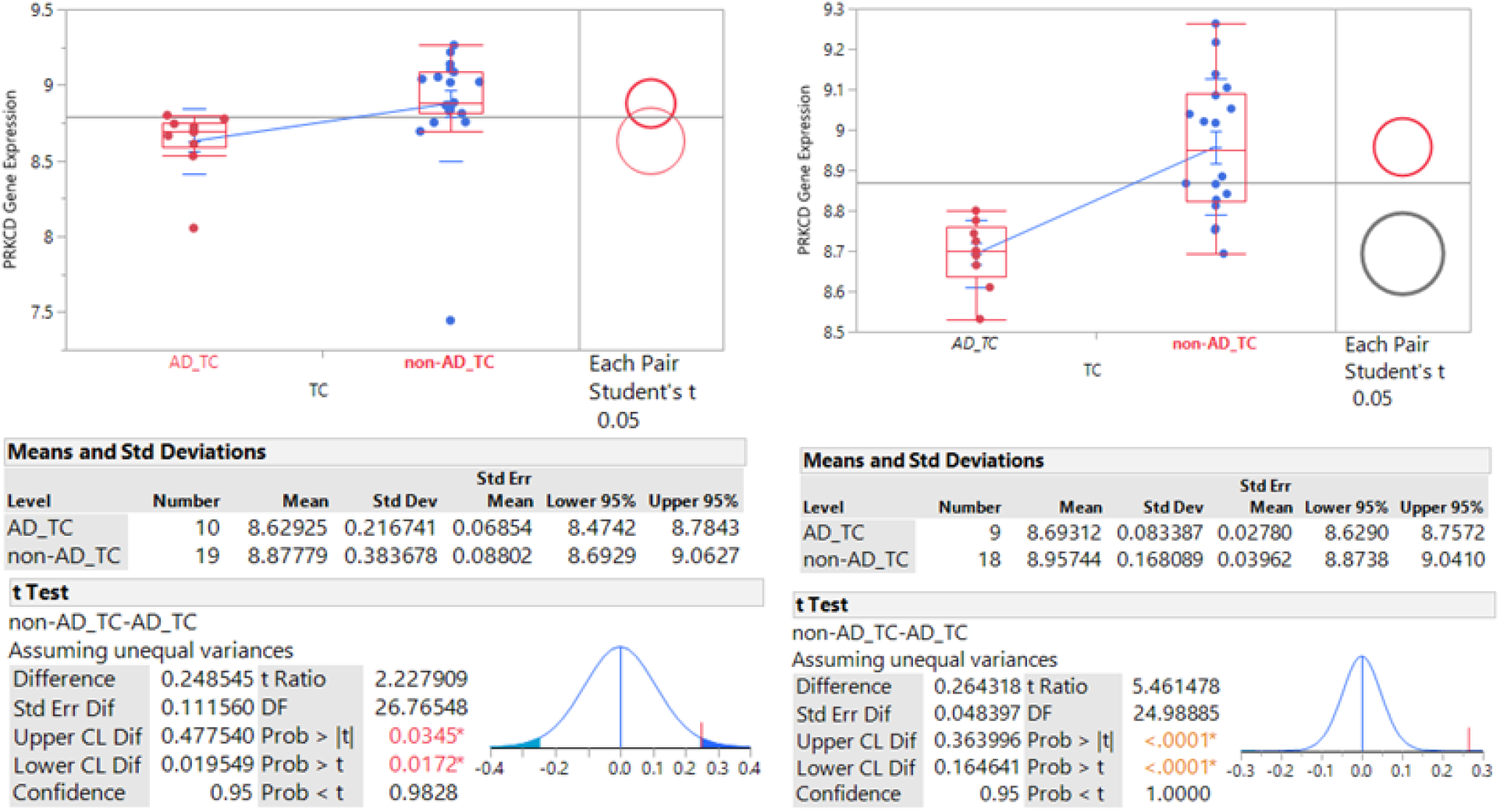
PRKCD gene expression in the TC a) on the left including all data points and b) on the right excluding outliers. Before modification of data, the data was already statistically significant, but after excluding outliers, the p-value was less than 0.0001.

Based on SOURCE, GRB2 is a growth factor receptor-bound protein 2, which has two SH3 domains that direct complex formation with proline-rich regions of other proteins, and also has SH2 domain that binds tyrosine phosphorylated sequences.

Based on SOURCE, PRKCD is protein kinase C delta, which is a part of the protein kinase C (PKC) family and is involved in B cell signaling and in the regulation of growth, apoptosis, and differentiation of a variety of cell types.

## Discussion

AD is a neurodegenerative disease that involves tau tangles and amyloid plaques in the brain. On the other hand, the blood and heart play a significant role in CVD development. There is a genetic predisposition in both diseases as well as similar risk factors. Analyzing the blood-brain relationship could be a potential path for treatment of AD by preventing CVD.

Two microarray datasets of AD and two microarray datasets of CVD, specifically myocardial infarction and heart failure, were analyzed to identify 73 differentially expressed genes (Appendix 1). All these genes were statistically significant, having a p-value less than 0.05, and were grouped into underexpressed (logFC<0) or overexpressed (logFC>0) genes. There were 31 common underexpressed and 42 common overexpressed genes in all 4 datasets. The genes that had the most protein-protein interactions were MAPK14, TLR2, HCK, GRB2, PRKCD, and PTPN6, signifying their importance in the relationship between AD and CVD.

MAPK14 (mitogen-activated protein kinase 14) had the most interactions of all the genes with a degree of 14. Another study identified MAPK14 as a possible Alzheimer’s therapeutic target [22] in regards to microglial activation and inflammation-induced synaptic toxicity reduction, which can help in clearing amyloid plaques. MAPK14 also mitigates defects in the autophagy-lysosomal system, which is a critical component of the pathogenesis of AD. Additionally, there has not been convincing evidence of brain developmental defects from neuronal deficiency of MAPK14, but research on humans will provide verification on using MAPK14 as a therapeutic target in AD. On the other hand with CVD, MAPK14 has been identified as a biomarker for cardioembolic stroke [23], which was not included in this study but still provides support for targeting MAPK14 in AD and CVD pathogenesis.

TLR2 (toll-like receptor 2) had the second most interactions of all the genes with a degree of 10. Another study identified TLR2 as a primary receptor for beta-amyloid to trigger neuroinflammatory activation. Liu et. al. concluded that TLR2 inhibition in microglia can be favorable in Alzheimer’s disease pathogenesis [24]. For CVDs, studies have reported toll-like receptors to play a role in heart failure (HF) and thus inhibition of TLRs can be beneficial in progression of HF [25]. TLR2 specifically contributes to activation of innate immunity in injured myocardium, which then contributes to myocardial inflammation, having a direct effect on HF.

The six genes of interest with the highest degrees were then analyzed in specific brain regions of AD and non-AD units. After conducting two-sample t-tests for differences in AD and non-AD mean, only two genes passed: GRB2 and PRKCD. The only common brain region in both genes that were statistically significant was the HI, which is expected since the HI plays a prominent role in memory. These two genes may indicate a relationship between the blood and brain as well as CVD and AD.

Previous studies have identified GRB2 as fundamental in atherosclerotic lesion formation [26], cardiac hypertrophy and fibrosis [27], all of which are in support of GRB2’s involvement in CVDs pathogenesis. Zhang et. al. hypothesizes that GRB2 promotes cardiac hypertrophy via a Gab1-PI3K-Akt pathway, which is highly expressed in heart tissue. Because GRB2 couples growth factor receptor activation to downstream mitogen-activated protein kinase cascades, GRB2 is linked with MAPK14, another gene of interest previously explored. In regards to AD, research has shown that GRB2 mediates trafficking of AbetaPP intracellular domain (AICD) “adaptor” protein that gets concentrated in neuronal cell bodies in AD patients [28].

In previous studies on PRKCD, it was identified that after an ischemic stroke, the altered protein kinase C (PKC) plays a role in blood brain barrier (BBB) disturbance and reperfusion damage. Furthermore, in people with obesity, PKC also plays a role in aortic contraction and adipocyte apoptosis. Following neural damage, PRKCD is increased, which aids in the formation and progression of Aβ disease like AD. Targeting PKCs pharmacologically could maintain the BBB and reduce ischemic infarct. PRKCD is heavily involved in mediating Aβ42 processing,which can result in cell lysis and the release of reactive oxygen species [29].

Genes involved in xylulose biosynthetic process, toll-like receptor TLR1:TLR2, and cellular response to triacyl bacterial lipopeptide were over 100 times enriched. The pathways, insulin effects increased synthesis of xylulose-5-phosphate,formyl peptide receptors bind formyl peptides and many other ligands, MyD88 deficiency (TLR2/4), pentose phosphate pathway, and the cellular components ficolin-1-rich granule and ficolin-1-rich granule lumen were significantly enriched as well. The available literature on these areas is limited, so future research could look at targeting these biological pathways.

### Limitations

Because of the lack of published datasets on AD and CVD patients, the size of the data pool is limited. The generalization of CVDs to heart attack and heart failure in this study may not be comprehensive. Future studies could focus on other types of CVDs such as coronary artery disease, cardiomyopathy, atherosclerosis, etc. Another potential area of limitation is the samples from the data used in this study aren’t consistent with the general population’s characteristics. For instance, in the dataset GSE36980, all samples came from Japan and the study is from the 1960s, so the updated datasets may be helpful for future more accurate study. The lack of distinction between female and male samples may be a caveat since the different sexes may yield slightly different gene expressions, which should be taken into account for treatment. The genes identified by this study were not compared to other neurodegenerative diseases, so it is not confirmed that these genes are unique only to AD.

### Future Directions

Instead of focusing on the peripheral blood and the brain, future studies could study specifically the connection between heart tissue and the brain for a more comprehensive analysis of the link between CVD and AD. Studying SNPs in the identified genes from this study could also help to further understand how genetic variants contribute to the association or development of both diseases. Future plans for this study could include a deeper study on the molecular structures of the genes of interest (GRB2, PRKCD). This could provide potential alternative paths of exploration for the development of medications to diminish the pathogenesis of AD.

## Conclusions

The underlying mechanism governing the association between AD and CVD may be related to GRB2 and PRKCD because both genes are differentially expressed in the brain and blood of AD and CVD patients; however, more research is required to validate that these genes can be used in the treatment or diagnosis of the diseases. Though these two genes may be of primary interest, the other 4 genes that had the most protein interactions in the gene network (MAPK14, TLR2, HCK, PTPN6) are notable. The 73 shared genes in all four datasets indicate commonality between AD and CVD as well. Other possible avenues for treatment of disease are in the upregulated pathways (insulin effects increased synthesis of xylulose-5-Phosphate, formyl peptide receptors bind formyl peptides and many other ligands, MyD88 deficiency (TLR2/4), pentose phosphate pathway) and biological processes (xylulose biosynthetic process, toll-like receptor TLR1:TLR2, and cellular response to triacyl bacterial lipopeptide).

## Data Availability

This study was using pre-existing gene expression data available freely from the NCBI Gene Expression Omnibus.

## Acknowledgments

I would like to thank Dr. Anna Delprato from the BioScience Project for supporting me in this project and teaching me the basic bioinformatics concepts used in this study.

## Appendix

| Underexpressed |  |  | Overexpressed |  |  |
| --- | --- | --- | --- | --- | --- |
| Gene Symbol |  |  | Gene Symbol |  |  |
| KLRB1 | CLK1 | COIL | GRN | ARAP3 | TLR2 |
| PSMB1 | YPEL5 | RABGGTB | LILRB3 | SP110 | MAPK14 |
| ATP5C1 | CGGBP1 | GTF3A | NCF4 | LILRA2 | ENO1 |
| NACA | PPP1R2 | CCNH | FPR2 | PTPN6 | SUOX |
| SSBP1 | HNRNPDL | MGAT4A | DCUN1D1 | NSFL1C | PLEC |
| DCTN6 | TMED10 | TRMO | RNF24 | TALDO1 | DUSP3 |
| UBE3A | NET1 | SLC25A36 | DUSP1 | TNFRSF1A | TMCO3 |
| WDR61 | ZNF277 | HSF2 | HSPA6 | RPS6KA1 | FKBP15 |
| EIF3M | ZNF451 | IDI1 | FPR1 | MCL1 | SPIDR |
| PSMD6 | API5 | GTPBP8 | TLR5 | HCK | ETS2 |
|  |  | AKT3 | CD14 | GNAI2 | GRB2 |
|  |  |  | PRKCD | WDR1 | DHPS |
|  |  |  | GNS | H2AFY | LILRA6 |
|  |  |  | TKT | SIRPA | PAFAH1B1 |

## Appendix 1.

Final gene list (total 73 genes) consisting of 31 underexpressed and 42 overexpressed genes that were shared among all 4 datasets for the blood analysis (GSE63060, GSE63061, GSE48060, and GSE9128)

## Appendix 2.

Link to STRING Network: https://string-db.org/cgi/network?taskId=bybBU0uqPsTj&sessionId=bqRPit3Lat8J

## References

[1] Kochanek KD, Murphy SL, Xu JQ, Arias E. (2019). Deaths: Final data for 2017. National Vital Statistics Reports, Vol 68 No 9. Hyattsville, MD: National Center for Health Statistics.

[2] Lyketsos, C. G., Carrillo, M. C., Ryan, J. M., Khachaturian, A. S., Trzepacz, P., Amatniek, J., Cedarbaum, J., Brashear, R., & Miller, D. S. (2011). Neuropsychiatric symptoms in Alzheimer’s disease. Alzheimer’s & Dementia, 7(5), 532–539. https://doi.org/10.1016/j.jalz.2011.05.2410

[3] Alzheimer’s Disease Fact Sheet. (2019). National Institute on Aging. https://www.nia.nih.gov/health/alzheimers-disease-fact-sheet

[4] Lin, Y.-T., Seo, J., Gao, F., Feldman, H. M., Wen, H.-L., Penney, J., Cam, H. P., Gjoneska, E., Raja, W. K., Cheng, J., Rueda, R., Kritskiy, O., Abdurrob, F., Peng, Z., Milo, B., Yu, C. J., Elmsaouri, S., Dey, D., Ko, T., … Tsai, L.-H. (2018). APOE4 Causes Widespread Molecular and Cellular Alterations Associated with Alzheimer’s Disease Phenotypes in Human iPSC-Derived Brain Cell Types. Neuron, 98(6), 1141-1154.e7. https://doi.org/10.1016/j.neuron.2018.05.008

[5] Liu, C.-C., Kanekiyo, T., Xu, H., & Bu, G. (2013). Apolipoprotein E and Alzheimer disease: risk, mechanisms and therapy. Nature Reviews Neurology, 9(2), 106–118. https://doi.org/10.1038/nrneurol.2012.263

[6] Dean, D. C., Jerskey, B. A., Chen, K., Protas, H., Thiyyagura, P., Roontiva, A., O’Muircheartaigh, J., Dirks, H., Waskiewicz, N., Lehman, K., Siniard, A. L., Turk, M. N., Hua, X., Madsen, S. K., Thompson, P. M., Fleisher, A. S., Huentelman, M. J., Deoni, S. C. L., & Reiman, E. M. (2014). Brain Differences in Infants at Differential Genetic Risk for Late-Onset Alzheimer Disease. JAMA Neurology, 71(1), 11. https://doi.org/10.1001/jamaneurol.2013.4544

[7] Heart Disease Facts | cdc.gov. (2020, September 8). Centers for Disease Control and Prevention. https://www.cdc.gov/heartdisease/facts.htm

[8] Lloyd-Jones, D. M., Nam, B.-H., D’Agostino, Sr R. B., Levy, D., Murabito, J. M., Wang, T. J., Wilson, P. W. F., & O’Donnell, C. J. (2004). Parental Cardiovascular Disease as a Risk Factor for Cardiovascular Disease in Middle-aged Adults. JAMA, 291(18), 2204. https://doi.org/10.1001/jama.291.18.2204

[9] Heart attack - Symptoms and causes. (2020, June 16). Mayo Clinic. https://www.mayoclinic.org/diseases-conditions/heart-attack/symptoms-causes/syc-20373106

[10] Heart failure - Symptoms and causes. (2020, May 29). Mayo Clinic. https://www.mayoclinic.org/diseases-conditions/heart-failure/symptoms-causes/syc-20373142

[11] Heart Failure | cdc.gov. (2020, September 8). Centers for Disease Control and Prevention. https://www.cdc.gov/heartdisease/heart_failure.htm#:%7E:text=About%206.2%20million%20adults%20in%20the%20United%20States%20have%20heart%20failure.

[12] Nabel, E. G. (2003). Cardiovascular Disease. New England Journal of Medicine, 349(1), 60–72. https://doi.org/10.1056/nejmra035098

[13] Yamada, Y., Izawa, H., Ichihara, S., Takatsu, F., Ishihara, H., Hirayama, H., Sone, T., Tanaka, M., & Yokota, M. (2002). Prediction of the Risk of Myocardial Infarction from Polymorphisms in Candidate Genes. New England Journal of Medicine, 347(24), 1916–1923. https://doi.org/10.1056/nejmoa021445

[14] Small, K. M., Wagoner, L. E., Levin, A. M., Kardia, S. L. R., & Liggett, S. B. (2002). Synergistic Polymorphisms of β1-and α2C-Adrenergic Receptors and the Risk of Congestive Heart Failure. New England Journal of Medicine, 347(15), 1135–1142. https://doi.org/10.1056/nejmoa020803

[15] White, L., Petrovitch, H., Hardman, J., Nelson, J., Davis, D. G., Ross, G. W., Masaki, K., Launer, L., & Markesbery, W. R. (2002). Cerebrovascular Pathology and Dementia in Autopsied Honolulu-Asia Aging Study Participants. Annals of the New York Academy of Sciences, 977(1), 9–23. https://doi.org/10.1111/j.1749-6632.2002.tb04794.x

[16] Newman, A. B., Fitzpatrick, A. L., Lopez, O., Jackson, S., Lyketsos, C., Jagust, W., Ives, D., DeKosky, S. T., & Kuller, L. H. (2005). Dementia and Alzheimer’s Disease Incidence in Relationship to Cardiovascular Disease in the Cardiovascular Health Study Cohort. Journal of the American Geriatrics Society, 53(7), 1101–1107. https://doi.org/10.1111/j.1532-5415.2005.53360.x

[17] Tini, G., Scagliola, R., Monacelli, F., La Malfa, G., Porto, I., Brunelli, C., & Rosa, G. M. (2020). Alzheimer’s Disease and Cardiovascular Disease: A Particular Association. Cardiology Research and Practice, 2020, 1–10. https://doi.org/10.1155/2020/2617970

[18] Stampfer, M. J. (2006). Cardiovascular disease and Alzheimer’s disease: common links. Journal of Internal Medicine, 260(3), 211–223. https://doi.org/10.1111/j.1365-2796.2006.01687.x

[19] Tublin, J. M., Adelstein, J. M., del Monte, F., Combs, C. K., & Wold, L. E. (2019). Getting to the Heart of Alzheimer Disease. Circulation Research, 124(1), 142–149. https://doi.org/10.1161/circresaha.118.313563

[20] The Hisayama Study. (2010). The Hisayama Study. https://www.hisayama.med.kyushu-u.ac.jp/en/

[21] NetworkAnalyzer Help. (2018). NetworkAnalyzer Online Help. https://med.bioinf.mpi-inf.mpg.de/netanalyzer/help/2.5/#:%7E:text=The%20topological%20coefficient%20is%20a,coefficient%20of%200%20(zero).

[22] Alam, J., & Scheper, W. (2016). Targeting neuronal MAPK14/p38α activity to modulate autophagy in the Alzheimer disease brain. Autophagy, 12(12), 2516–2520. https://doi.org/10.1080/15548627.2016.1238555

[23] Li, Z., Xu, L., & Wang, Q. (2020). Integrative Analysis of MAPK14 as a Potential Biomarker for Cardioembolic Stroke. BioMed Research International, 2020, 1–15. https://doi.org/10.1155/2020/9502820

[24] Liu, S., Liu, Y., Hao, W., Wolf, L., Kiliaan, A. J., Penke, B., Rübe, C. E., Walter, J., Heneka, M. T., Hartmann, T., Menger, M. D., & Fassbender, K. (2011). TLR2 Is a Primary Receptor for Alzheimer’s Amyloid β Peptide To Trigger Neuroinflammatory Activation. The Journal of Immunology, 188(3), 1098–1107. https://doi.org/10.4049/jimmunol.1101121

[25] Yu, L., & Feng, Z. (2018). The Role of Toll-Like Receptor Signaling in the Progression of Heart Failure. Mediators of Inflammation, 2018, 1–11. https://doi.org/10.1155/2018/9874109

[26] Proctor, B. M., Ren, J., Chen, Z., Schneider, J. G., Coleman, T., Lupu, T. S., Semenkovich, C. F., & Muslin, A. J. (2007). Grb2 Is Required for Atherosclerotic Lesion Formation. Arteriosclerosis, Thrombosis, and Vascular Biology, 27(6), 1361–1367. https://doi.org/10.1161/atvbaha.106.134007

[27] Zhang, S., Weinheimer, C., Courtois, M., Kovacs, A., Zhang, C. E., Cheng, A. M., Wang, Y., & Muslin, A. J. (2003). The role of the Grb2–p38 MAPK signaling pathway in cardiac hypertrophy and fibrosis. Journal of Clinical Investigation, 111(6), 833–841. https://doi.org/10.1172/jci16290

[28] Raychaudhuri, M., & Mukhopadhyay, D. (2010). Grb2-Mediated Alteration in the Trafficking of AβPP: Insights from Grb2-AICD Interaction. Journal of Alzheimer’s Disease, 20(1), 275–292. https://doi.org/10.3233/jad-2010-1371

[29] Lucke-Wold, B. P., Turner, R. C., Logsdon, A. F., Simpkins, J. W., Alkon, D. L., Smith, K. E., Chen, Y.-W., Tan, Z., Huber, J. D., & Rosen, C. L. (2014). Common Mechanisms of Alzheimer’s Disease and Ischemic Stroke: The Role of Protein Kinase C in the Progression of Age-Related Neurodegeneration. Journal of Alzheimer’s Disease, 43(3), 711–724. https://doi.org/10.3233/jad-141422

